# AncestryDNA COVID-19 Host Genetic Study Identifies Three Novel Loci

**DOI:** 10.1101/2020.10.06.20205864

**Authors:** Genevieve H.L. Roberts, Danny S. Park, Marie V. Coignet, Shannon R. McCurdy, Spencer C. Knight, Raghavendran Partha, Brooke Rhead, Miao Zhang, Nathan Berkowitz, AncestryDNA Science Team, Asher K. Haug Baltzell, Harendra Guturu, Ahna R. Girshick, Kristin A. Rand, Eurie L. Hong, Catherine A. Ball

**Author notes:** These authors contributed equally to this work (Genevieve H.L. Roberts, Danny S. Park). **Corresponding Author:** Catherine Ball.

## Abstract

Human infection with SARS-CoV-2, the causative agent of COVID-19, leads to a remarkably diverse spectrum of outcomes, ranging from asymptomatic to fatal. Recent reports suggest that both clinical and genetic risk factors may contribute to COVID-19 susceptibility and severity. To investigate genetic risk factors, we collected over 500,000 COVID-19 survey responses between April and May 2020 with accompanying genetic data from the AncestryDNA database. We conducted sex-stratified and meta-analyzed genome-wide association studies (GWAS) for COVID-19 susceptibility (positive nasopharyngeal swab test, *n*_*cases*_=2,407) and severity (hospitalization, *n*_*cases*_=250). The severity GWAS replicated associations with severe COVID-19 near *ABO* and *SLC6A20* (*P*<0.05). Furthermore, we identified three novel loci with *P*<5×10^−8^. The strongest association was near *IVNS1ABP*, a gene involved in influenza virus replication^1^, and was associated only in males. The other two novel loci harbor genes with established roles in viral replication or immunity: *SRRM1* and the immunoglobulin lambda locus. We thus present new evidence that host genetic variation likely contributes to COVID-19 outcomes and demonstrate the value of large-scale, self-reported data as a mechanism to rapidly address a health crisis.

## Introduction

Novel severe acute respiratory syndrome coronavirus 2 (SARS-CoV-2), the causative agent of COVID-19, precipitated a pandemic with >21 million cases and >760,000 deaths worldwide as of August 2020.^2^ Outcomes of SARS-CoV-2 infection in the United States are diverse; most infections result in mild illness that can be managed at home, yet ∼14% of cases are hospitalized and ∼5% are fatal.^3^ Epidemiological studies have identified clinical risk factors for severe COVID-19 that include common health conditions such as hypertension, diabetes, obesity, older age, and male sex.^4,5^ Reports of higher susceptibility to^6,7,8^ and severity of^9^ SARS-CoV infections in men could suggest important biological differences in immune response to SARS-CoV-2 in men relative to women.^10^

In addition to clinical risk factors, emerging evidence suggests that host genetic variation may contribute to COVID-19 susceptibility and severity. Ellinghaus *et al*. conducted a genome-wide association study (GWAS) of COVID-19 cases with respiratory failure and identified two loci that achieved genome-wide significance: one signal on chromosome (chr) 9 near the *ABO* gene, which determines blood type, and one signal on chr 3 near a cluster of genes with known immune function including *SLC6A20, CXCR6, CCR1, CCR2*, and *CCR9*.^11^ Additionally, a small whole-exome sequencing study identified *TLR7*, an X-chromosome gene involved in interferon signal induction, in four male patients with severe COVID-19.^12^ To validate rapidly emerging results from new studies such as the Ellinghaus study, further investigation in independent datasets is needed. Furthermore, investigation in larger datasets with increased statistical power may detect additional, novel host genetic variation relevant to COVID-19 susceptibility and severity.

To replicate and discover non-genetic^6^ and genetic associations with COVID-19 outcomes, we engaged AncestryDNA members who have consented to research in the United States, with 18 million total individuals in the global network. On April 22, 2020, we released a 54-question COVID-19 survey intended to assess exposure, risk factors, symptomatology, and demographic information that had been previously identified as associated with COVID-19 susceptibility and severity in the evolving pandemic. In under two months, over 500,000 COVID-19 survey responses were collected with a 95% survey completion rate.

From these self-reported outcomes, we constructed two phenotypes: one intended to assess susceptibility, in which individuals who reported a positive COVID-19 test were compared to those who reported a negative test (referred to throughout as “susceptibility”) and one intended to assess severity, in which who were hospitalized with COVID-19 were compared to COVID-19 positive individuals who were not hospitalized (referred to throughout as “hospitalization”). To identify novel genetic determinants of these outcomes, we conducted a GWAS for each phenotype in a cohort of European-descent individuals. Sex-stratified GWAS were performed to investigate potential biological sex-driven differences in immune response to SARS-CoV-2 infection, and the GWAS results were meta-analyzed to maximize statistical power for variant discovery.

## Results

### COVID-19 survey results and cohort demographics

To validate the COVID-19 survey, we examined how representative our cohort is with respect to the United States population both in terms of COVID-19 infection status and demographics. The COVID-19 survey presented to AncestryDNA customers who consented to research has between 39 and 54 questions, depending on reported COVID-19 test result **(Supplementary Figure 1 and Supplementary Table 1)**. The first survey question assesses testing status, and in total, 3,733, or 13.4% of respondents who were tested, reported a positive test result **(Supplementary Table 2)** – a proportion comparable to the national cumulative positivity rate of 12% during a similar collection period (March 1 - May 30, 2020).^13^ Of respondents that reported a positive COVID-19 test, 375 (11%) reported hospitalization, comparable to a U.S. Centers for Disease Control and Prevention (CDC) report of a 14% hospitalization rate (**Supplementary Table 3**).^3^

Extended demographic data presented in **Supplementary Table 3** show that the proportion of COVID-19 survey respondents with European ancestry is largely consistent with the U.S. population: roughly 75% of respondents were classified as having European ancestry, comparable to 73% of the U.S. general population that self-identify as “white” in the United States Census Bureau’s 2018 American Community Survey.^14^ In the COVID-19 survey, 65% of respondents were female, and the United States Census Bureau’s 2018 American Community Survey reports a female population of 50.8%. This is consistent with previous literature reporting gender differences in willingness to participate in survey completion.^15^ The median age of COVID-19 survey respondents was 56, notably higher than the national median age^14^ of 38; however, a minimum age of 18 was required to participate in the study.

The survey data were filtered to select a final GWAS cohort to analyze (filtering steps summarized in **Supplementary Figure 2**). **Table 1** presents the final GWAS cohort sample sizes for the susceptibility and severity phenotypes. In total, 2,417 cases (862 male, 1,555 female) and 14,933 controls (4,472 male, 10,461 female) were used in the susceptibility GWAS and 250 cases (105 male, 145 female) and 1,967 controls (679 male, 1,288 female) were used in the hospitalization GWAS.

**Table 1:** Total number of cases and controls for the susceptibility and hospitalization GWAS

|  | Susceptibility |  | Hospitalization |  |
| --- | --- | --- | --- | --- |
|  | Cases<br>COVID-19<br>Positive | Controls<br>COVID-19<br>Negative | Cases<br>COVID-19 Cases<br>Reporting<br>Hospitalization | Controls<br>COVID-19 Cases<br>Reporting NO<br>Hospitalization |
| All* | 2,417 | 14,933 | 250 | 1,967 |
| Male* | 862 | 4,472 | 105 | 679 |
| Female* | 1,555 | 10,461 | 145 | 1,288 |
| Age (median)* | 52 | 55 | 58 | 50 |

\*All individuals in the GWAS cohort are of European ancestry. A summary of exclusion criteria is presented in Supplementary Figure 2.

### Replication of *ABO* and *SLC6A20* COVID-19 severity associations

Ellinghaus *et al*. (2020) performed a GWAS consisting of 1,980 Spanish and Italian COVID-19 cases with respiratory failure and 1,805 controls, most of whom had not been tested for COVID-19. This study identified two loci that achieved genome-wide significance: one signal on chr 3 represented by rs11385942 near the *SLC6A20* gene and one signal on chr 9 represented by rs657152 near the *ABO* gene.^11^

We assessed replication of the lead SNPs at the two loci identified by Ellinghaus *et al*.^11^ within our hospitalization GWAS. Although the phenotypes are not the same, both respiratory failure and hospitalization assess aspects of COVID-19 infection severity. As summarized in **Table 2**, we observed nominal replication of the lead SNPs at these two loci in our hospitalization analysis at both the *ABO* locus (*P*=0.022) and the *SLC6A20* locus (*P*=0.020). For both loci, consistent risk alleles and directions of effect were observed, but with generally smaller odds ratios (ORs) than those reported by Ellinghaus and colleagues. We did not observe significant associations at either locus in the susceptibility analysis **(Supplementary Table 4)**.

**Table 2:**
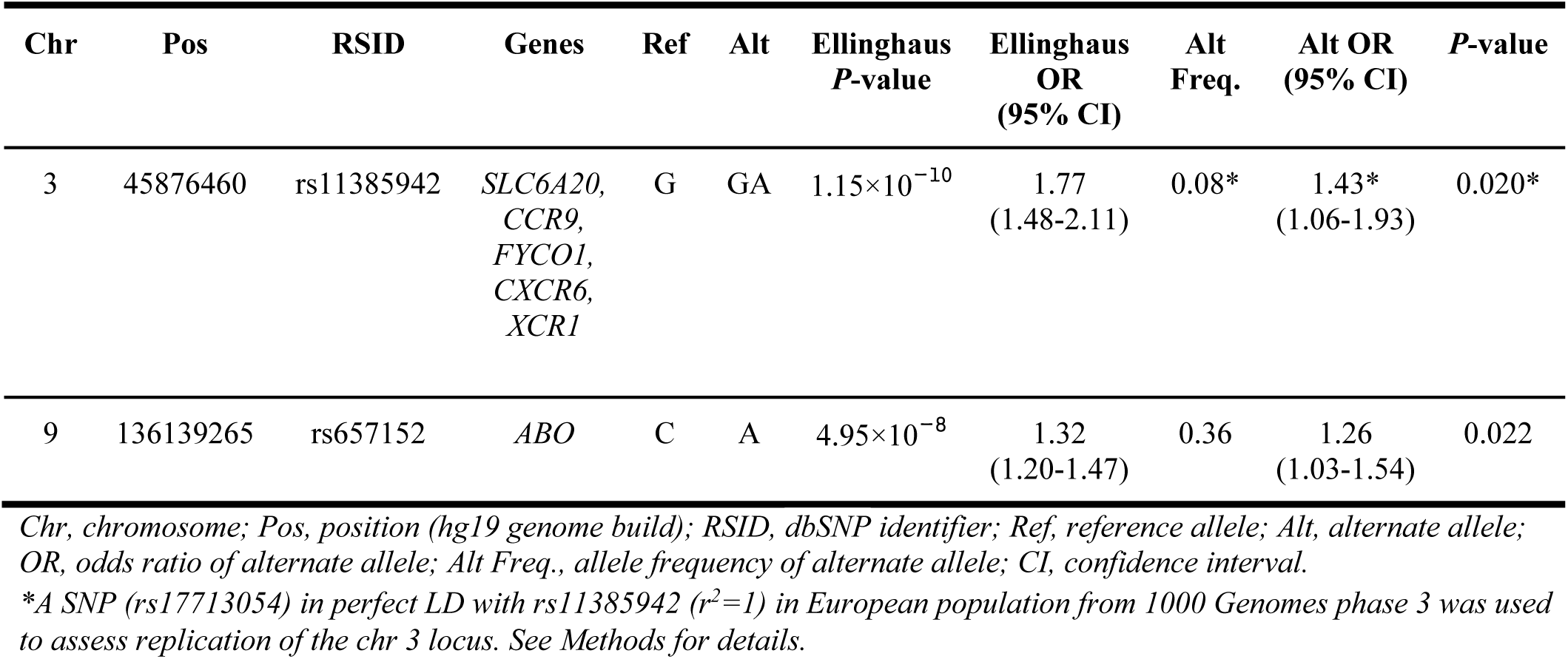
Replication of severe respiratory failure loci from Ellinghaus *et. al*. (2020)^11^ with hospitalization phenotype

### Novel associations with COVID-19 susceptibility and severity

We conducted a sex-stratified GWAS adjusted for orthogonal age, orthogonal age^2^, array version, and PC1-12 (**Supplementary Table 5)**, to investigate possible differences in genetic associations with COVID-19 outcomes in males and females and meta-analyzed the resulting summary statistics to maximize statistical power. For the susceptibility phenotype, the male GWAS, female GWAS, and meta-analysis had genomic inflation factors of 1.00, 1.01, and 1.00, respectively **(Supplementary Figure 3)**. The hospitalization GWAS had genomic inflation factors of 1.04, 1.01, and 0.99 for males, females, and the meta-analysis, respectively **(Supplementary Figure 4)**. In total, three novel loci surpassed the genome-wide significance threshold of 5×10^−8^ in at least one study: two separate loci on chr 1 and one locus on chr 22 **(Table 3 and Figure 1, Supplementary Figures 5 and 6)**.

**Table 3:**
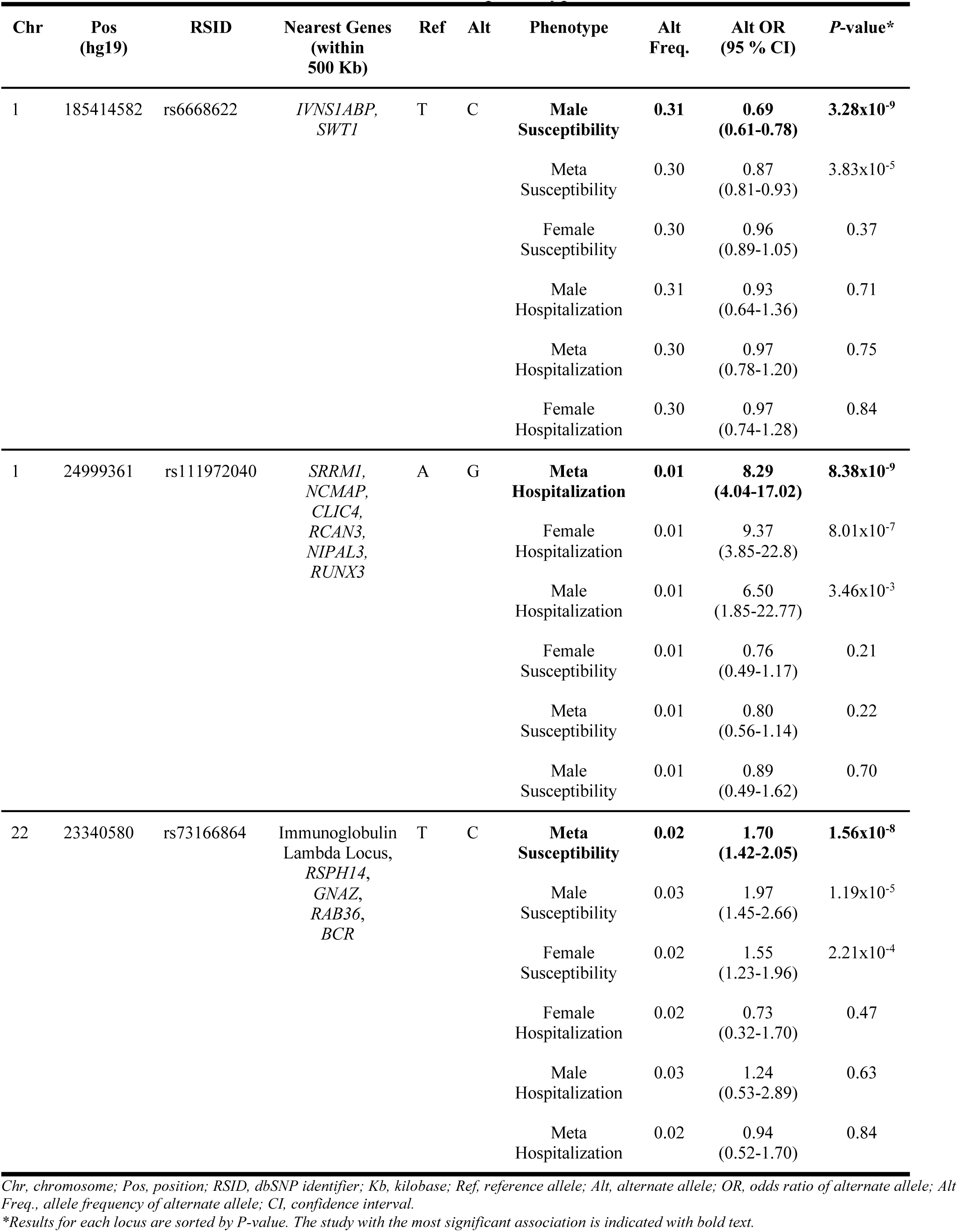
Novel loci with *P*<5×10^−8^ for one or more phenotypes

**Figure 1:**
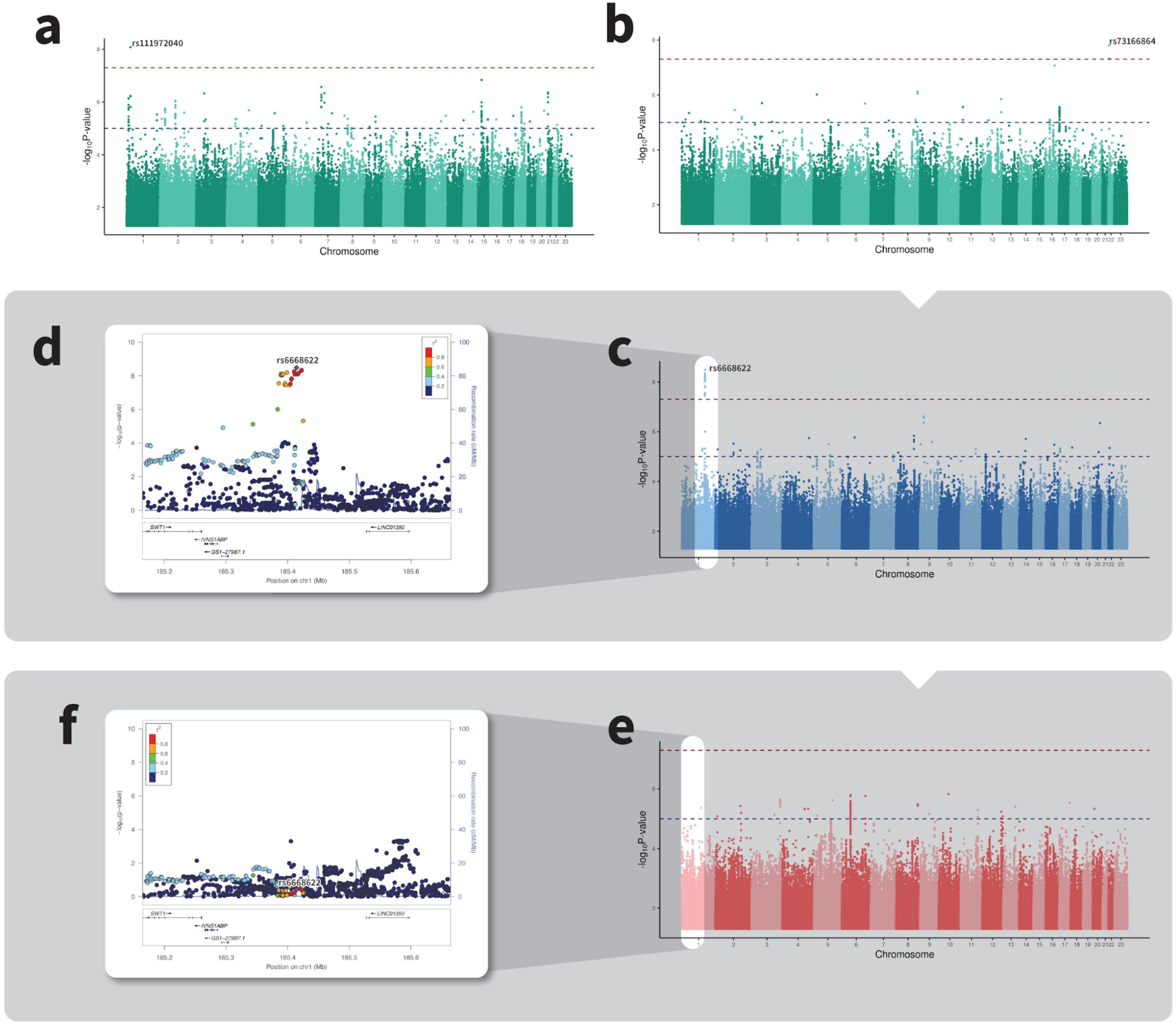
Manhattan plots with horizontal red dashed line representing genome-wide significance (*P*=5×10^−8^) and horizontal blue dashed line representing suggestive significance (*P*=1×10^−5^) for the **(a)** hospitalization meta-analysis **(b)** susceptibility meta-analysis **(c)** susceptibility GWAS in males only (blue), highlighting the association peak in the *IVNS1ABP* region represented by rs6668622 **(d)** LocusZoom plot indexed on rs6668622 in male-only susceptibility GWAS **(e)** susceptibility GWAS in females only (red), highlighting the absence of an association peak in the *IVNS1ABP* region represented by rs6668622 **(f)** LocusZoom plot indexed on rs6668622 in female-only susceptibility GWAS.

In the susceptibility analysis, the most significant association was represented by lead SNP rs6668622, with the association present in males only (*P*=3.28×10^−9^; OR=0.69) **(Figure 1c-d)** and absent in females (*P*=0.37; OR=0.96) **(Figure 1e-f)**. In the sex-combined meta-analysis, the rs6668622 association did not reach genome-wide significance (*P*=3.83×10^−5^; OR=0.87). Consistent with the differential association observed in men and women, there is significant heterogeneity of effect (*I*^*2*^) for rs6668622 between the male and the female studies (*I*^*2*^=94; *P*=1.6×10^−5^; **Supplementary Table 6**). This signal is intergenic and the nearest protein-coding genes to rs6668622 are *IVNS1ABP* (∼150Kb) and *SWT1* (∼288Kb) **(Supplementary Figure 7)**.

We identified an additional locus associated with susceptibility on chr 22, represented by lead SNP rs73166864. This locus achieved genome-wide significance in the meta-analysis (*P*=1.56×10^−8^; OR=1.70); the effect sizes were not significantly different (*I*^*2*^*=*1.6; *P*=0.20; **Supplementary Table 6**) in the male (*P*=1.19×10^−5^; OR=1.97) and female (*P*=2.21×10^−4^; OR=1.55) analyses. The variant rs73166864 is intergenic and is within 500kb of the immunoglobulin lambda locus^16^, which encodes proteins used to construct the light chains of antibodies. The nearest protein-coding genes to rs73166864 are *IGLL5, GNAZ, RSPH14, RAB36* and *BCR* **(Supplementary Figure 8)**.

In the hospitalization meta-analysis, a locus on chr 1 surpassed genome-wide significance. The lead SNP, rs111972040, is uncommon with a MAF of approximately 1%, but with a large estimated effect size in the hospitalization meta-analysis (*P*=8.38×10^−9^; OR=8.29). Although rs111972040 did not achieve genome-wide significance in either sex-stratified GWAS, the estimated ORs in the male (*P*=3.46×10^−3^; OR=6.50) and female (*P*=8.01×10^−7^; OR=9.37) studies were large and there was no significant heterogeneity of effect between males and females (*I*^*2*^=0; *P*=0.63; **Supplementary Table 6**). The variant rs111972040 is a non-coding transcript variant in the gene *SRRM1*, and *NCMAP, CLIC4, RCAN3, NIPAL3*, and *RUNX3* are all within 500kb **(Supplementary Figure 9)**.

To assess whether clinical risk factors other than age and sex had an effect on these associated loci, we additionally adjusted for other associated risk factors including body mass index (BMI) and having one or more pre-existing health conditions (asthma, bone marrow transplant, cancer, cardiovascular disease, kidney disease, chronic obstructive pulmonary disease (COPD), diabetes, hypertension, organ transplant, autoimmune disease, immunodeficiency, or lung conditions). For all three novel lead SNPs, the estimated effect sizes remained relatively consistent, though the *P*-value dropped below genome-wide significance in the hospitalization analysis, likely in part due to the small sample size and the additional decrease in sample size for these extended analyses **(Supplementary Table 7)**. Both loci associated with susceptibility remained genome-wide significant even after adjusting for these additional clinical risk factors.

### Heritability of COVID-19 susceptibility and severity

We estimated narrow-sense heritability (*h*^*2*^) – defined as the proportion of phenotypic variation due to additive genetic factors^17^ – using the linear mixed model approach (GCTA-GREML)^18^ and autosomal genome-wide imputed variants. The underlying assumption of this linear mixed model approach is that the included common variants all contribute equally small effects to heritability.^19^

To our knowledge, these are the first estimates of heritability for COVID-19 susceptibility and severity (hospitalization) based on empirical genetic similarity, though one twin-based study estimated heritability of COVID-19 symptoms.^20^ As shown in **Supplementary Table 8**, for susceptibility, the estimated liability heritability was *h*^*2*^ =0.00 (standard error [SE]=0.07). The estimated liability heritability for hospitalization was *h*^*2*^=0.14 (SE=0.58); the large standard error is partly a result of the small hospitalization sample size.

## Discussion

To identify genetic determinants of COVID-19 susceptibility and severity, we conducted GWAS of self-reported COVID-19 outcomes in a population of survey respondents with European ancestry. To explore possible differences in biological response to SARS-CoV-2 infection, we analyzed both susceptibility and severity outcomes via sex-stratified GWAS and sex-combined meta-analyses. In total, three novel loci surpassed genome-wide significance in one or more study, with lead SNPs rs6668622 (male susceptibility *P*=3.2×10^−9^), rs73166864 (susceptibility *P*=1.56×10^−8^), and rs111972040 (hospitalization *P*=8.38×10^−9^) near *IVNS1ABP*, the immunoglobulin lambda locus, and *SRRM1*, respectively.

The most significantly associated SNP in any study was rs6668622 with the susceptibility outcome. The nearest gene to rs6668622 is *IVNS1ABP*, which encodes Influenza Virus NS1A Binding Protein. This protein is known to bind with influenza virulence factor NS1 and this interaction appears to promote influenza viral gene expression.^21^ The variant rs6668622 is a known, strong expression quantitative trait locus (eQTL) in lung tissue for *IVNS1ABP*^22,23^, suggesting that risk variation might impact mRNA abundance of *IVNS1ABP*. Strikingly, haploinsufficiency of *IVNS1ABP* appears to associate with primary immunodeficiency^24^, suggesting *IVNS1ABP* may play a role in cellular response to other pathogens besides influenza. It is unclear why this association is only present only in males, though it may provide a clue as to why males appear to be at higher risk of COVID-19 infection, hospitalization, and mortality.^7-12^ We speculate that sex hormones or behavioral differences might trigger generally different cellular responses to SARS-Cov-2 infection in men and in women^10^, and one such difference may involve differential expression of *IVNS1ABP*.

Another locus, represented by the intergenic lead SNP rs73166864, was associated with the susceptibility outcome with similar effects in men and women. This signal is ∼75Kb away from the immunoglobulin lambda locus, a region that undergoes somatic recombination in B-cells and encodes proteins used to construct the antigen-binding light chain region of antibodies.^16^ It is unclear what the functional consequence of this intergenic variation might be, but proximity to such an important region for antibody generation is intriguing.

The final locus, represented by the lead SNP rs111972040, was the only genome-wide significant association with the hospitalization outcome. The variant rs111972040 is a non-coding transcript variant in the gene *SRRM1*, which encodes Serine and Arginine Repetitive Matrix 1. A related gene, *SRRM2*, was implicated in human immunodeficiency virus (HIV) splicing and replication.^25^ Based on this, we hypothesize that *SRRM1* may likewise play an important role in splicing SARS-Cov-2 viral genes.

In addition to identifying novel associations, the severity GWAS replicated findings from a previous COVID-19 respiratory failure GWAS^11^ that identified two loci: the blood type *ABO* gene and a cluster of immune genes near *SLC6A20*. We observed consistent directions of effect at both loci, but the replication *P*-values were nominal, and we observed smaller estimated ORs; however, these observations are not surprising. The original study considered a more severe outcome (respiratory failure) than our analogous severity study (hospitalization). Furthermore, we included only 250 cases that were hospitalized in our study relative to the 1,980 cases with respiratory failure in the Ellinghaus study^11^, thus the nominal *P*-values in this study may simply reflect lower statistical power for our severity analysis. Lastly, the winner’s curse suggests that overestimated effect sizes are expected in the discovery study.^26^

We estimated heritability for both outcomes to assess the contribution of common genetic variation to COVID-19 susceptibility and severity. The liability heritability estimates were generally small: *h*^*2*^=0.00 (SE=0.07) for susceptibility and *h*^*2*^=0.14 (SE=0.58) for hospitalization. However, for hospitalization, the sample sizes were small for this type of analysis and standard errors were correspondingly high. A key assumption underlying the heritability estimates is that many small effects were distributed across the entire genome^19^. Thus, the low heritability estimates might not simply reflect low genetic contribution, but rather could suggest a genetic architecture involving a limited number of high-effect variants or a large contribution by uncommon and rare variants.

A key limitation of our data is that COVID-19 cases who suffered very severe or fatal infections are less likely to participate in our survey, which consequently results in undersampling cases with severe outcomes. We also restricted to individuals of European ancestry due to small sample sizes in other genetic ancestry cohorts for the susceptibility and severity outcomes in this early phase of COVID-19 survey data collection. As the COVID-19 survey cohort grows, future analyses will focus on increased ancestral diversity to increase generalizability. Finally, we lack an independent replication cohort for our novel findings and will rely on future ascertainment of additional survey respondents and COVID-19 GWAS consortia^27^ efforts to determine whether our findings are reproducible.

In summary, we collected over 500,000 self-reported COVID-19 outcomes in under two months and conducted one of the largest genetic studies of infection susceptibility and severity to date, thus demonstrating the value of large-scale self-reported data as a mechanism to rapidly address a serious health crisis. We identified three novel loci, all of which harbor genes with established roles in viral replication or immunity, and one of which may provide insight into why men appear to be differently affected by COVID-19 than women. We thus add to growing evidence that host genetic variation contributes to COVID-19 susceptibility and severity and suggest identification of such genetic risk factors may provide profound insight into pathogenesis of the novel coronavirus.

## Methods

### Ethics statement

All data for this research project was from subjects who provided prior informed consent to participate in AncestryDNA’s Human Diversity Project, as reviewed and approved by our external institutional review board, Advarra (formerly Quorum). All data was de-identified prior to use.

### Study population

Self-reported COVID-19 outcomes were collected through the Personal Discoveries Project^®^, a survey platform available to AncestryDNA customers via the web and mobile applications. The COVID-19 survey ranged from 39-54 questions, depending on the initial COVID-19 test result reported. **Supplementary Figure 1** describes the flow of the topics assessed in each section of the survey. The full questions and possible responses for the two questions used as primary outcomes in this study are presented in **Supplementary Table 1**. Analyses presented here were performed with data collected between April 22-May 28, 2020.

To participate in the COVID-19 survey, participants must meet the following criteria: they must be 18 years of age or older, a resident of the United States, be an existing AncestryDNA customer who has consented to participate in research^28^, and be able to complete a short survey. The survey is designed to assess self-reported COVID-19 positivity and severity, as well as susceptibility and known risk factors including community exposure and known contacts with individuals diagnosed with COVID-19.

### Phenotype definitions

Two phenotypes were assessed: one for susceptibility and one for severity of COVID-19. Cases for the COVID-19 susceptibility phenotype were defined as individuals who responded to the question, “Have you been swab tested for COVID-19, commonly referred to as coronavirus?” as “Yes, and was positive”. Responders who answered “Yes, and was negative” were defined as controls for the susceptibility study. Cases for the severity phenotype were defined as individuals who reported testing positive for COVID-19 and responded to the question, “Were you hospitalized due to these symptoms?” as “Yes”. Severity controls reported testing positive for COVID-19, but reported no hospitalization related to COVID-19.

### Genotyping

Genotyping and quality control procedures have been previously described elsewhere.^28^ Briefly, customer genotype data for this study were generated using an Illumina genotyping array with approximately 730,000 SNPs and processed either with Illumina or with Quest/Athena Diagnostics. To ensure quality of each dataset, a sample passes a number of quality control (QC) checks, which includes identifying duplicate samples, removing individuals with a per-sample call rate <98%, and identifying discrepancies between reported sex and genetically inferred sex. Samples that pass all quality-control tests proceed to the analysis pipeline; samples that fail one or more tests must be recollected or manually cleared for analysis by lab technicians. Array markers with per-variant call rate <0.98 and array markers that had overall allele frequency differences of >0.10 between any two array versions were additionally removed prior to downstream analyses.

### Selecting a European ancestry association cohort

As a measure to control population stratification, we selected individuals with estimated European ancestry for inclusion in the GWAS. To determine these definitions, a proprietary algorithm was used to estimate continental admixture proportions for all COVID-19 survey respondents.^29^ Briefly, this algorithm uses a hidden Markov model to estimate unphased diploid ancestry across the genome by comparing haplotype structure to a reference panel. The reference panel consists of a combination of AncestryDNA customers and publicly available datasets and is designed to reflect global diversity. From our total cohort of 506,743 individuals who participated in the COVID-19 survey, 379,383 (74.9%) individuals with estimated European ancestry were retained **(Supplementary Table 3 and Supplementary Figure 10)**.

### Removal of related individuals

AncestryDNA’s identity-by-descent inference algorithm^30^ was used to estimate the relationship between pairs of individuals. Pairs with estimated separation of fewer than four meioses were considered close relatives. For all close relative pairs, one individual was randomly selected for exclusion from our study. We excluded 1,741 individuals from the susceptibility analysis and 223 from the severity analysis due to relatedness.

### Calculation of principal components to control residual population structure

After selecting unrelated individuals with European ancestry as described above, genetic PCs were calculated to include in the association studies to control residual population structure and were computed using FlashPCA 2.0.^31^ Input genotypes were linkage disequilibrum (LD)-pruned using PLINK 1.9 command `--indep-pairwise 100 5 0.2 --maf 0.05 --geno 0.001`.

### Imputation

Samples were imputed to the Haplotype Reference Consortium (HRC) reference panel^32^ version 1.1, which consists of 27,165 total individuals and 36 million variants. The HRC reference panel does not include indels; consequently, indels are not present in the results of our analyses. We determined best-guess haplotypes with Eagle version 2.4.1^33^ and performed imputation with Minimac4 version 1.0.1.^34^ We used 1,117,080 unique variants as input and 8,049,082 imputed variants were retained in the final data set. For these variants, we conservatively restricted our analyses to variants minor allele frequency (MAF)>0.01 and Minimac4 R^2^>0.30 using genotype dosage probabilities for all variants regardless of whether they were originally genotyped.

### Statistical analyses

We used the COVID-19 Host Genetics Initiative (HGI) analysis plan version 1 to guide our analyses.^35^ A key recommendation in this plan is to analyze males and females separately when possible; therefore, we conducted four separate GWAS in total: susceptibility in males, susceptibility in females, hospitalization in males, and hospitalization in females. Sex was determined from genotype data. For each GWAS, a fixed-effects logistic regression model was implemented with PLINK2.0 with either the susceptibility or severity phenotype as the primary outcome and imputed genotype dosage value as the primary predictor. The following were included as fixed-effect covariates: PCs 1-12 (described above), array platform, orthogonal age, and orthogonal age^2^. Orthogonal polynomials were used to eliminate collinearity between age and age^2^ and were calculated in R version 3.6.0 with base function `poly(age, degree=2)`. We additionally used PLINK2.0 to remove variants with Minimac4 imputation quality R^2^<0.3 or with MAF<0.01. The following PLINK2.0 flags were used for each analysis:

~~~
  --vcf [input imputed VCF] dosage=DS
  --psam [file that provides sex information]
  --covar [covariates file]
  --covar-name PC1, PC2, PC3, PC4, PC5, PC6, PC7, PC8, PC9, PC10, PC11, PC12,
  orthogonal_age, orthogonal_age^2^, platform
  --covar-variance-standardize
  --extract-if-info R2 >= 0.3
  --freq
  --glm hide-covar
  --keep [list of unrelated Europeans]
  --keep-females OR keep-males
  --maf 0.01
  --pheno [phenotype file]
  --pheno-name [phenotype column name]
~~~

For each phenotype, summary statistics for males and females were combined using a fixed-effects inverse-variance weighted meta-analysis, implemented with METAL^36^ (version released 25 March 2011). Unless otherwise noted, all variant ORs are adjusted for the 15 covariates described above. For all individual GWAS and meta-analyses, we considered the European genome-wide significance threshold^37^ of two-tailed *P*<5×10^−8^ to represent a significant association. For lead SNPs at loci surpassing *P*<5×10^−8^, we performed extended analyses in which we additionally adjusted for BMI and self-reported affliction with one or more of any of the following health conditions: asthma, bone marrow transplant, cancer, cardiovascular disease, kidney disease, COPD, diabetes, hypertension, organ failure requiring transplant, autoimmune disease, immunodeficiency, and/or “other” lung condition.

### Replication

To our knowledge, the only published GWAS of COVID-19 outcomes to date was performed by Ellinghaus *et al*.^11^ We compared *P*-values and OR estimates from our analyses to the two lead variants reported by Ellinghaus: rs657152, representing a region on chr 9 near *ABO*; and rs11385942, representing a region on chr 3 near *SLC6A20*. The variant rs11385942 is a small indel and indels are not present in the HRC reference panel; we therefore examined the association with rs11385942 using rs17713054, a SNP in perfect LD (R^2^=1, D’=1) in a European population from LDpair.^38^ The allele rs17713054-A corresponds to rs11385942-AG.

### Heritability

To estimate heritability of COVID-19 phenotypes, we calculated phenotypic variance explained by a genetic relatedness matrix (GRM) in the European GWAS cohort. To calculate the GRM, we first performed LD-pruning of autosomal, imputed, high-quality (Minimac4 R^2^>0.3) best-guess genotypes with PLINK-1.9 command `--indep-pairwise 100 5 0.2 --maf 0.05 –geno 0.001 --chr 1-22`, resulting in a total of 224,096 variants. From these variants, GCTA version 1.91.5 beta2 was used to estimate a GRM.^18^ We subset the resulting matrix to unrelated individuals (`--grm-cutoff 0.025`) and individuals with diagonal elements between 0.95-1.05, resulting in a final GRM consisting of 18,415 subjects. These 18,415 individuals are candidates for the two heritability analyses.

Next, we performed single component heritability estimation for each phenotype by fitting a linear mixed model (GCTA-GREML)^18^. Each model included PCs 1-12, array platform, sex inferred from genotype data, orthogonal age, and orthogonal age^2^ as fixed effects. For each phenotype, the model also included a random genetic effect with the covariance given by a GRM calculated with the intersection of the 18,415 heritability individuals and those with non-missing phenotypes using the same 224,096 variants described above.

The observed heritability was estimated using the GCTA command --grm [phenotype-specific GRM] --reml --pheno [phenotype file] --covar [genetic sex, platform] --qcovar [PCs 1-12, orthogonal_age, orthogonal_age^2^]. The liability heritability was estimated from the observed heritability using the standard transformation^39^ and the sample prevalence using the GCTA command above with `--prevalence [sample prevalence].`

## Data Availability

Additional supplementary data files may be shared with qualified researchers on a case by case basis. Please reach out to.

## Footnotes

## Acknowledgements

We thank our AncestryDNA customers who made this study possible by contributing information about their experience with COVID-19 through our survey. Without them, this work would not be possible. We would like to thank Zach Bass, Robert Dowling, Disha Akarte, Swapnil Sneham, Sean Enright and the entire Cyborg team for their tireless work in the release and continued support of the COVID-19 survey. We would like to acknowledge Chris Trainor and David Serventi for their work on the Figures. We additionally thank Mark Daly and the COVID-19 Host Genetics Consortium for advice and preliminary feedback.

## Author contributions

GHLR and DSP contributed equally to the manuscript. GHLR and DSP wrote the manuscript with support from MVC and SRM. GHLR and DSP designed and conducted genome-wide association studies, GHLR and DSP interpreted results with support from MVC. SCK, RP, and BR helped with additional analyses and interpretation. MZ, HG, SCK, NB, AHB, and DSP performed genotype imputation and data management. MVC and KAR designed the COVID-19 survey questionnaire and MVC assessed concordance of phenotype prevalence with the U.S. population. SRM conducted heritability analyses. ARG, AHB, and HG facilitated forward progression of the manuscript and provided input and guidance. The AncestryDNA Science Team contributed to additional work, allowing for the completion of the COVID-19 research and manuscript. KAR led the COVID-19 research and data teams. KAR, ELH, and CAB provided project guidance. All authors have contributed to and reviewed the final manuscript.

## Competing interests

The authors declare competing financial interests: authors affiliated with AncestryDNA are employed by Ancestry and may have equity in Ancestry.

## AncestryDNA Science Team

Yambazi Banda, Ke Bi, Robert Burton, Marjan Champine, Ross Curtis, Karen Delgado, Abby Drokhlyansky, Ashley Elrick, Cat Foo, Michael Gaddis, Jialiang Gu, Heather Harris, Shannon Hateley, Shea King, Christine Maldonado, Evan McCartney-Melstad, Alexandra McFarland, Patty Miller, Luong Nguyen, Keith Noto, Milos Pavlovic, Jingwen Pei, Jenna Petersen, Scott Pew, Chodon Sass, Josh Schraiber, Alisa Sedghifar, Andrey Smelter, Sarah South, Barry Starr, David Turissini, Cecily Vaughn, Yong Wang

## Notes

### Funding Statement

All work was supported and funded by AncestryDNA

### Author Declarations

All data for this research project was from subjects who provided prior informed consent to participate in AncestryDNAs Human Diversity Project, as reviewed and approved by our external institutional review board, Advarra (formerly Quorum). All data was de-identified prior to use.

