## Supplementary Tables and Figures for "AncestryDNA COVID-19 Host Genetic Study Identifies Three Novel Loci"

**AncestryDNA Science Team:**

Yambazi Banda
Ke Bi
Robert Burton
Marjan Champine
Ross Curtis
Karen Delgado
Abby Drokhlyansky
Ashley Elrick
Cat Foo
Michael Gaddis
Jialiang Gu
Heather Harris
Shannon Hateley
Shea King
Christine Maldonado
Evan McCartney-Melstad
Alexandra McFarland
Patty Miller
Luong Nguyen
Keith Noto
Milos Pavlovic
Jingwen Pei
Jenna Petersen
Scott Pew
Chodon Sass
Josh Schraiber
Alisa Sedghifar
Andrey Smelter
Sarah South
Barry Starr
David Turissini
Cecily Vaughn
Yong Wang

### TABLE OF CONTENTS


SUPPLEMENTARY TABLES 2

**Supplementary Table 1:** Survey questions used to create hospitalization and susceptibility phenotypes 2

**Supplementary Table 2:** COVID-19 testing status cohort-wide 3

**Supplementary Table 3:** Extended demographic information for COVID-19 study cohort 4

**Supplementary Table 4:** Extended replication results for Ellinghaus *et al.* (2020) 6

**Supplementary Table 5:** Association of GWAS PCs with COVID-19 hospitalization and susceptibility phenotypes 7

**Supplementary Table 6:** Extended association results for novel loci with $P$<5x10$^{-8}$ 8

**Supplementary Table 7:** Extended covariate adjustment results for novel loci with GWAS $P$<5x10$^{-8}$ 9

**Supplementary Table 8:** Extended genetic heritability estimates 10

SUPPLEMENTARY FIGURES 11

**Supplementary Figure 1:** Survey flow 11

**Supplementary Figure 2:** Filtering individuals for inclusion in GWAS 12

**Supplementary Figure 3:** QQplots for susceptibility GWAS 13

**Supplementary Figure 4:** QQplots for hospitalization GWAS 14

**Supplementary Figure 5:** Manhattan plots for susceptibility GWAS 15

**Supplementary Figure 6:** Manhattan plots for hospitalization GWAS 16

**Supplementary Figure 7:** LocusZoom plots for *IVNS1ABP* locus 17

**Supplementary Figure 8:** LocusZoom plots for immunoglobulin lambda locus 18

**Supplementary Figure 9:** LocusZoom plots for *SRRM1* locus 19

**Supplementary Figure 10:** Principal components plot of continental ancestry groups 20

### SUPPLEMENTARY TABLES

**Supplementary Table 1:** Survey questions used to create hospitalization and susceptibility phenotypes

| Question | Answer Options | | | | |
|---|---|---|---|---|---|
| *Have you been swab tested for COVID-19, commonly referred to as coronavirus?* | Yes, and was positive | Yes, and was negative | Yes, and my results are pending | No, but I have had flu-like symptoms with a fever at some point since the beginning of February 2020 | No, and I have not had flu-like symptoms at some point since the beginning of February 2020 |
| *Were you hospitalized due to these symptoms?* | Yes | No | Not sure | | |

**Supplementary Table 2:** COVID-19 testing status cohort-wide

| COVID-19 Status | Count (%) |
| --- | --- |
| Yes, and was positive | 3,733 (1%) |
| Yes, and was negative | 21,422 (4%) |
| Yes, and my results are pending | 2,672 (1%) |
| No, but I have had flu-like symptoms with a fever at some point since the beginning of February | 65,970 (13%) |
| No, and I have not had flu-like symptoms at some point since the beginning of February | 412,946 (81%) |

**Supplementary Table 3:** Extended demographic information for COVID-19 study cohort

| | COVID-19 Nasopharyngeal Swab Test Positive<br><br>*n* = 3,733 | COVID-19 Nasopharyngeal Swab Test Negative<br><br>*n* = 21,422 | Full Ancestry COVID-19 Cohort<br><br>*n* = 506,743 |
|---|---|---|---|
| **Median Age** | 49 | 53 | 56 |
| **Self-reported Gender** | | | |
| Female | 2,097 (56%) | 14,374 (67%) | 328,262 (65%) |
| Male | 1,138 (31%) | 6,111 (29%) | 156,329 (31%) |
| Non-binary, other, or prefer not to state | 498 (13%) | 937 (4%) | 22,913 (4%) |
| **Genetic Sex** | | | |
| Female | 2,399 (64%) | 14,907 (70%) | 341,780 (67%) |
| Male | 1,328 (36%) | 6,472 (30%) | 164,773 (33%) |
| **Genetic Ancestry Continental Grouping**[1] | | | |
| Admixed African-European ancestry | 217 (6%) | 908 (4%) | 14,759 (3%) |
| European ancestry | 2,419 (65%) | 14,970 (70%) | 378,868 (75%) |
| Admixed Amerindian ancestry | 409 (11%) | 1,743 (8%) | 32,749 (7%) |
| Other ancestry | 688 (18%) | 3,801 (18%) | 80,367 (15%) |
| **Risk Factors** | | | |
| Asthma | 566 (13%) | 4,174 (14%) | 66,002 (10%) |
| Cardiovascular disease | 161 (4%) | 1,312 (5%) | 26,565 (4%) |
| Diabetes | 319 (7%) | 2,179 (7%) | 47,526 (8%) |
| Hypertension | 703 (16%) | 5,062 (17%) | 116,626 (18%) |
| Autoimmune disease | 348 (8%) | 2,736 (9%) | 47,381 (7%) |
| Rare health conditions[2] | 331 (9%) | 3,366 (16%) | 50,278 (10%) |
| At least one of above conditions | 1,559 (46%) | 11,466 (55%) | 231,047 (47%) |
| "Other" condition | 204 (5%) | 1,395 (5%) | 25,417 (4%) |
| Not sure | 75 (2%) | 470 (2%) | 10,892 (2%) |
| None | 1,654 (38%) | 8,428 (29%) | 242,074 (38%) |
| **Smoking Status** | | | |
| Ever | 1,279 (40%) | 9,352 (46%) | 213,148 (44%) |
| Never | 1,946 (60%) | 11,064 (54%) | 269,490 (56%) |
| Current | 156 (12%) | 1,740 (19%) | 35,575 (17%) |
| Median pack years for smokers | 10 | 12 | 13 |
| **Median BMI** | 28.7 | 28.7 | 28.2 |
| **Influenza** | | | |
| Vaccine (yes) | 2,173 (64%) | 14,835 (71%) | 321,558 (65%) |
| Diagnosis (yes) | 156 (5%) | 899 (4%) | 10,496 (2%) |
| **Symptoms** | 3,167 (90%) | - | 3,863 (64%)[4] |
| **Hospitalization** | 375 (11%) | - | 1,203 (2%)[4] |
| Median duration, days (sd) | 5 (8) | - | 3 (7)[4] |

| | | | |
|---|---|---|---|
| Oxygen[3] | 166 (45%) | - | 312 (26%)[4] |
| Ventilation[3] | 35 (9%) | - | 64 (5%)[4] |

*sd, standard deviation*
*[1]Genetic ancestry grouping definitions: Admixed African-European ancestry includes 100% African ancestry; Admixed East Asian-European ancestry also includes 100% East Asian ancestry; Admixed Amerindian ancestry also includes 100% Amerindian ancestry. Supplementary Figure 10 shows principal components plots of ancestry groupings*
*[2]Rare health conditions had one or more cells with fewer than 100 individuals and includes: cancer, kidney disease, chronic obstructive pulmonary disease, lung condition, immunodeficiency, bone marrow transplant, and organ transplant.*
*[3]Percentages represent % of total hospitalized*
*[4]This includes COVID-19+, COVID-19 test pending, and individuals who were not tested but felt sick*

#### **Supplementary Table 4:** Extended replication results for Ellinghaus *et al.* (2020)

| Locus | Ellinghaus[1] Lead SNP RSID | LD Tag SNP RSID[2] | LD with with Ellinghaus Lead SNP | Ref | Alt | Imp. $R^2$ | Case Freq | Ctrl Freq | *β* (SE) | *P*-value | Dir | Het $I^2$ | Het. *P*-value |
|---|---|---|---|---|---|---|---|---|---|---|---|---|---|
| ***SLC6A20 Locus*** | | | | **G** | **A** | 1 | | | | | | | |
| Meta-analysis | **rs11385942** | **rs17713054** | **D'=1 $R^2$=1** | | | | N/A | N/A | 0.36 (0.15) | 0.02 | ++ | 0 | 0.65 |
| Males | | | | | | | 0.12 | 0.09 | 0.29 (0.26) | 0.27 | | | |
| Females | | | | | | | 0.12 | 0.09 | 0.39 (0.19) | 0.05 | | | |
| ***ABO Locus*** | | | | | | | | | | | | | |
| Meta-analysis | **rs657152** | **N/A** | **N/A** | **C** | **A** | 1 | N/A | N/A | 0.23 (0.10) | 0.02 | ++ | 13.5 | 0.28 |
| Males | | | | | | | 0.44 | 0.37 | 0.41 (0.17) | 0.01 | | | |
| Females | | | | | | | 0.4 | 0.37 | 0.16 (0.13) | 0.22 | | | |

*Chr, chromosome; Pos, position (hg19 genome build); RSID, dbSNP identifier; Ref, reference allele; Alt, alternate allele; Imp $R^2$; MINIMAC4 Imputation $R^2$; Freq, allele frequency of alternate allele; Ctrl, control; β, logistic regression effect estimate for the LD Tag SNP; Dir, Direction of effect for alternate allele in male-only GWAS GWAS and female-only GWAS; Het $I^2$, heterogeneity $I^2$ statistic; Het P-value, P-value for heterogeneity statistic.*

*[1]Lead SNP from Ellinghaus, D., et al. Genomewide association study of severe Covid-19 with respiratory failure. N Engl J Med. doi: 10.1056/NEJMoa2020283 (2020).*

*[2]If the lead variant was not present in our study, an LD Tag was used. D' and $R^2$ in this column represent LD statistics between the lead variant and the LD tag variant, as calculated in the European population from 1000 Genomes phase 3*

**Supplementary Table 5:** Association of GWAS PCs with COVID-19 hospitalization and susceptibility phenotypes

| | **Logistic Regression *β* (SE)[1]** | | | | **Logistic Regression *P*-value[1]** | | | |
|---|---|---|---|---|---|---|---|---|
| | **Hospitalization** | | **Susceptibility** | | **Hospitalization** | | **Susceptibility** | |
| | **Male** | **Female** | **Male** | **Female** | **Male** | **Female** | **Male** | **Female** |
| **PC1** | -2.22 (2.05) | 0.46 (2.17) | -3.98 (0.78) | -4.95 (0.67) | 0.279 | 0.831 | 2.03E-7 | 1.07E-13 |
| **PC2** | -8.09 (5.18) | -2.47 (4.90) | -2.35 (1.93) | -1.06 (1.47) | 0.118 | 0.613 | 0.206 | 0.471 |
| **PC3** | 7.82 (5.77) | -4.74 (4.82) | -2.73 (1.99) | -5.22 (1.55) | 0.175 | 0.325 | 0.155 | 7.59E-4 |
| **PC4** | -0.28 (8.67) | -3.10 (7.28) | 15.5 (3.02) | 13.12 (2.23) | 0.974 | 0.670 | 2.98E-7 | 4.25E-9 |
| **PC5** | -13.09 (13.02) | -13.70 (10.11) | -3.14 (4.33) | -5.68 (3.09) | 0.315 | 0.175 | 0.466 | 0.066 |
| **PC6** | 2.99 (12.77) | -4.93 (9.751) | 6.81 (4.22) | 0.90 (3.07) | 0.815 | 0.612 | 0.088 | 0.768 |
| **PC7** | 13.22 (13.40) | -0.05 (10.82) | 0.883 (4.39) | -0.42 (3.13) | 0.324 | 0.996 | 0.846 | 0.893 |
| **PC8** | -18.15 (13.26) | 7.09 (10.21) | 1.00 (4.25) | 0.24 (3.11) | 0.171 | 0.487 | 0.863 | 0.938 |
| **PC9** | -0.63 (13.43) | 3.36 (10.50) | 1.04 (4.37) | -0.12 (3.10) | 0.962 | 0.749 | 0.861 | 0.969 |
| **PC10** | 9.35 (12.99) | 1.84 (10.18) | -3.29 (4.35) | -0.66 (3.13) | 0.472 | 0.856 | 0.438 | 0.834 |
| **PC11** | -15.48 (12.84) | 16.58 (10.05) | 7.62 (4.29) | 0.64 (3.12) | 0.228 | 0.099 | 0.079 | 0.838 |
| **PC12** | 13.60 (12.81) | -7.15 (10.55) | -0.359 (4.27) | 0.10 (3.11) | 0.288 | 0.498 | 0.963 | 0.975 |
| **PC13[2]** | -10.15 (12.93) | -0.41 (10.54) | 3.23 (4.34) | 0.75 (3.16) | 0.432 | 0.968 | 0.449 | 0.813 |
| **PC14[2]** | 9.59 (12.77) | -4.71 (10.51) | -0.348 (4.32) | -2.71 (3.16) | 0.453 | 0.654 | 0.903 | 0.392 |
| **PC15[2]** | 20.68 (13.16) | 10.81 (10.63) | 3.76 (4.34) | 0.06 (3.15) | 0.116 | 0.309 | 0.37 | 0.984 |
| **PC16[2]** | -6.55 (13.07) | 6.58 (10.52) | 7.27 (4.38) | -0.50 (3.15) | 0.616 | 0.531 | 0.095 | 0.873 |
| **PC17[2]** | 12.34 (13.28) | 4.96 (10.28) | -2.42 (4.35) | -2.16 (3.18) | 0.353 | 0.629 | 0.524 | 0.498 |
| **PC18[2]** | 21.00 (13.45) | 8.32 (11.16) | 0.007 (4.41) | 0.15 (3.19) | 0.118 | 0.456 | 0.942 | 0.962 |
| **PC19[2]** | 17.89 (12.56) | -5.23 (10.56) | 1.03 (4.47) | -1.56 (3.16) | 0.154 | 0.620 | 0.819 | 0.621 |
| **PC20[2]** | -4.93 (13.05) | -0.91 (10.55) | 6.05 (4.38) | 2.53 (3.17) | 0.706 | 0.931 | 0.198 | 0.424 |

*β, logistic regression effect estimate; SE, standard error*

[1]*PC associations are adjusted for array platform, orthogonal age, and orthogonal age*$^2$

[2]*PCs 13-20 were not included in analyses. No PCs beyond PC13 were significantly associated (P<0.05) with either outcome.*

**Supplementary Table 6:** Extended association results for novel loci with $P<5x10^{-8}$

| | | Imputed | | | | | | | | Unimputed[1] | | | |
|---|---|---|---|---|---|---|---|---|---|---|---|---|---|
| | Lead RSID | Imp $R^2$ | Case Freq | Ctrl Freq | $\beta$ (SE) | P-value | Dir | Het $I^2$ | Het. P-Value | Lead SNP | LD with lead Imputed SNP[2] | SNP $\beta$ (SE) | P-value |
| ***IVNS1ABP* Locus/Susceptibility** | | | | | | | | | | | | | |
| Meta-analysis | **rs6668622** | 0.99 | NA | NA | -0.14 (0.04) | 3.8E-05 | -- | 94 | 1.6E-5 | rs10911736 | $R^2$=0.99 D'=1 | -0.14 (0.03) | 5.65E-05 |
| Males* | | | .242 | .317 | -0.37 (0.06) | 3.3E-09 | | | | | | -0.36 (0.06) | 5.41E-09 |
| Females | | | .289 | .302 | -0.04 (0.04) | 0.37 | | | | | | -0.03 (0.04) | 0.38 |
| ***SRRM1* Locus/Hospitalization** | | | | | | | | | | | | | |
| Meta-analysis* | **rs111972040** | 0.79 | NA | NA | 2.12 (0.37) | 8.4E-9 | ++ | 0 | 0.63 | rs4649014 | $R^2$=0.03 D'=1 | 0.35 (0.12) | 0.0034 |
| Males | | | .034 | .006 | 1.87 (0.64) | 0.0035 | | | | | | 0.45 (0.19) | 0.02 |
| Females | | | .039 | .007 | 2.24 (0.45) | 8.1E-7 | | | | | | 0.30 (0.15) | 0.059 |
| **Immunoglobulin Lambda Locus/Susceptibility** | | | | | | | | | | | | | |
| Meta-analysis* | **rs73166864** | 0.91 | NA | NA | 0.53 (0.09) | 1.6E-8 | ++ | 1.6 | 0.20 | rs4820538 | $R^2$=0.03 D'=0.82 | 0.12 (0.05) | 0.00913 |
| Males | | | .038 | .022 | 0.43 (0.15) | 1.2E-5 | | | | | | 0.19 (0.08) | 0.0226 |
| Females | | | .032 | .022 | 0.68 (0.12) | 0.00022 | | | | | | 0.09 (0.06) | 0.12 |

*Lead RSID, dbSNP identifier of the most associated variant at the locus; Imp $R^2$; MINIMAC4 Imputation $R^2$; Freq, allele frequency of alternate allele; Ctrl, control; β, logistic regression effect estimate; Dir, Direction of effect for alternate allele in male-only GWAS and female-only GWAS; Het $I^2$, heterogeneity $I^2$ statistic; Het P, P-value for heterogeneity statistic.*

**Denotes the study with the most significant association at the indicated locus*

*[1]The lead variant at each locus that was directly typed on a genotyping array*

*[2]D' and $R^2$ in this column represent LD statistics between the lead imputed variant and the lead unimputed (directly typed) variant, as calculated in the European population from 1000 Genomes phase 3*

**Supplementary Table 7:** Extended covariate adjustment results for novel loci with GWAS $P<5x10^{-8}$

| | ***IVNS1ABP* locus represented by rs6668622 with male susceptibility outcome** | | ***SRRM1* locus represented by rs111972040 with meta[1] hospitalization outcome** | | **Immunoglobulin lambda locus represented by rs73166864 with meta[1] susceptibility outcome** | |
|---|---|---|---|---|---|---|
| | SNP $\beta$ (SE) | SNP P-value | SNP $\beta$ (SE) | SNP P-value | SNP $\beta$ (SE) | SNP P-value |
| Standard Model[2] | 1.999 (0.062) | $3.29x10^{-9}$ | -2.115 (0.37) | $1.10x10^{-8}$ | 0.527 (0.094) | $2.17x10^{-8}$ |
| Standard Model + BMI[3] | 1.994 (0.067) | $3.54x10^{-8}$ | -1.903 (0.39) | $1.19\ x10^{-6}$ | 0.556 (0.10) | $3.43x10^{-8}$ |
| Standard Model + Any Health Condition[4] | 1.991 (0.065) | $1.22x10^{-8}$ | -1.931 (0.37) | $2.36x10^{-7}$ | 0.546 (0.098) | $2.45x10^{-8}$ |
| Standard Model + BMI[3] + Any Health Condition[4] | 1.994 (0.067) | $3.70x10^{-8}$ | -1.742 (0.39) | $1.10x10^{-5}$ | 0.555 (0.10) | $3.82x10^{-8}$ |

*BMI, body mass index*

[1]*A logistic regression model was fit in males and females separately and subsequently meta-analyzed with inverse variance weighting.*

[2]*The "Standard Model" was the model applied in sex-stratified GWAS analyses: logit(phenotype) ~ PC1:PC12 + orthogonal age + orthogonal age$^2$ + array platform*

[3]*Subjects that skipped the height and weight questions and outliers with reported BMI > 6 standard deviations above the mean or BMI < 2 standard deviations below the mean by sex were excluded from analyses that included BMI, resulting in 23 (9%) cases and 158 (8%) controls dropped for hospitalization and 375 (16%) cases and 830 (6%) controls dropped for susceptibility, thus, an increase in P-value is expected for models that include BMI.*

[4]*Any Health Condition refers to any of the 12 health conditions included in Supplementary Table 3, coded as "1" for any subject that reported having one or more of the 12 health conditions and "0" for all others.*

**Supplementary Table 8:** Extended genetic heritability estimates

| | **Susceptibility** | **Hospitalization** |
|---|---|---|
| *n* Total[1] | 16,622 | 2,090 |
| *n* Cases | 2,283 | 228 |
| *n* Controls | 14,339 | 1,862 |
| Case Prevalence[2] | 0.14 | 0.11 |
| Estimated $h^2$ on Observed Scale (SE)[3] | $1x10^{-6}$ (0.03) | 0.05 (0.21) |
| Estimated $h^2$ on Liability Scale (SE)[4] | $2x10^{-6}$ (0.07) | 0.14 (0.58) |

*SE, standard error; $h^2$, heritability*

[1]*The heritability "n Total", "n Cases", and "n Controls" are smaller than for GWAS due to additional relatedness filtering (see Methods)*

[2]*Case prevalence is used to transform heritability estimates from the observed to the liability scale*

[3]*The observed heritability, $h^2$, is the proportion of variance explained by the genetic relatedness matrix in a linear mixed model with outcome values $\in\{0,1\}$*

[3]*Observed heritability transformed to the liability scale*

### SUPPLEMENTARY FIGURES

**Supplementary Figure 1:** Survey flow

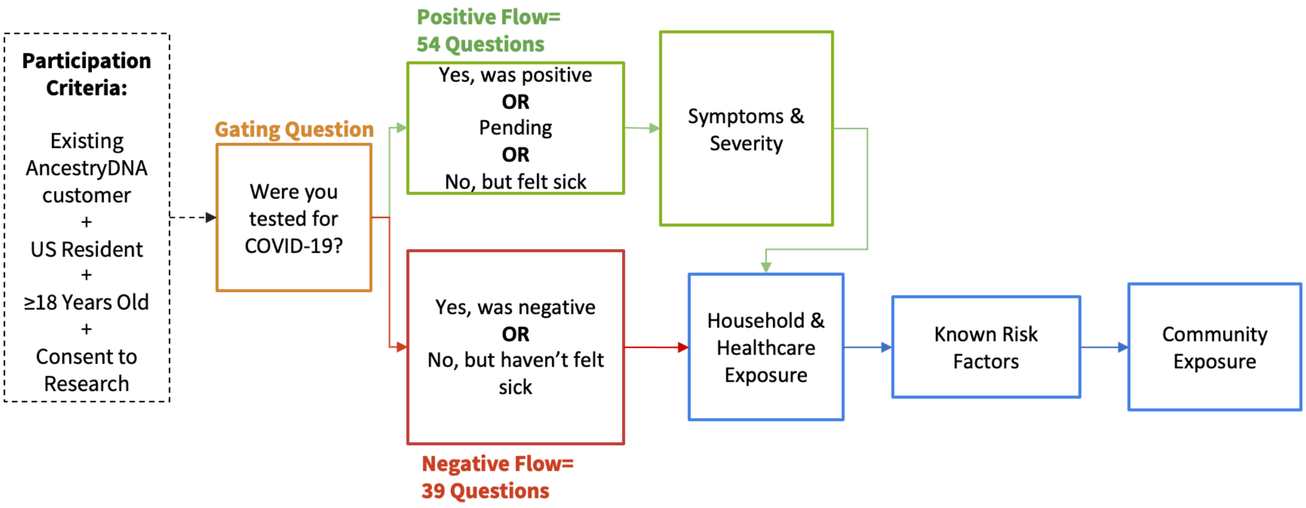


*Topical schema showing the flow and logic of the Ancestry COVID-19 survey. The gating question in Supplementary Table 1 assesses whether the respondent has been tested for COVID-19 then shuttles respondents into two different flows: one with symptom, hospitalization and complication assessment, and one that does not include those questions. Answers that prompt the detailed sickness assessment are: positive COVID-19 nasal swab test, tested but with results pending, and not tested but have had flu-like symptoms with a fever at some point since the beginning of February 2020. All survey participants answer questions related to potential exposures and suspected risk factors for COVID-19 susceptibility and severity.*

**Supplementary Figure 2:** Filtering individuals for inclusion in GWAS

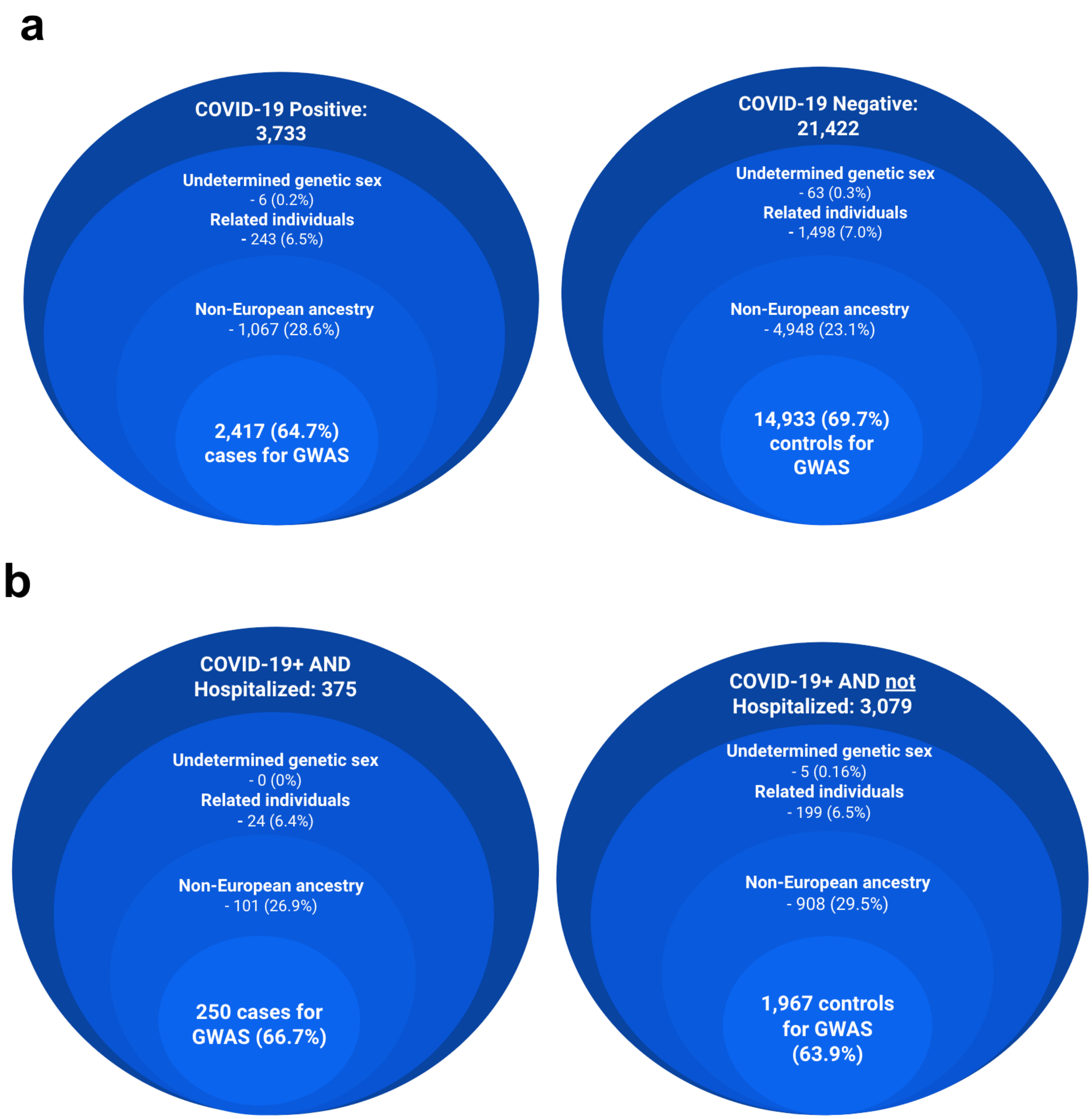


*Funnel schemas of filtering steps to finalized phenotypes.* ***(a)*** *Susceptibility phenotype begins with 3,733 COVID-19 positive (+) individuals and 21,422 COVID-19 negative (-) individuals; after excluding those with missing information and relatedness and filtering on genetic ancestry, 2,417 cases and 14,933 controls in the susceptibility GWAS.* ***(b)*** *The hospitalization phenotype begins with 375 people hospitalized and that tested positive for COVID-19 and 3,079 people that were not hospitalized and tested postitive for COVID-19; after excluding those with missing information and relatedness and filtering on genetic ancestry of 250 cases and 1,967 controls post filtering and exclusions due to missingness and relatedness.*

**Supplementary Figure 3:** QQplots for susceptibility GWAS

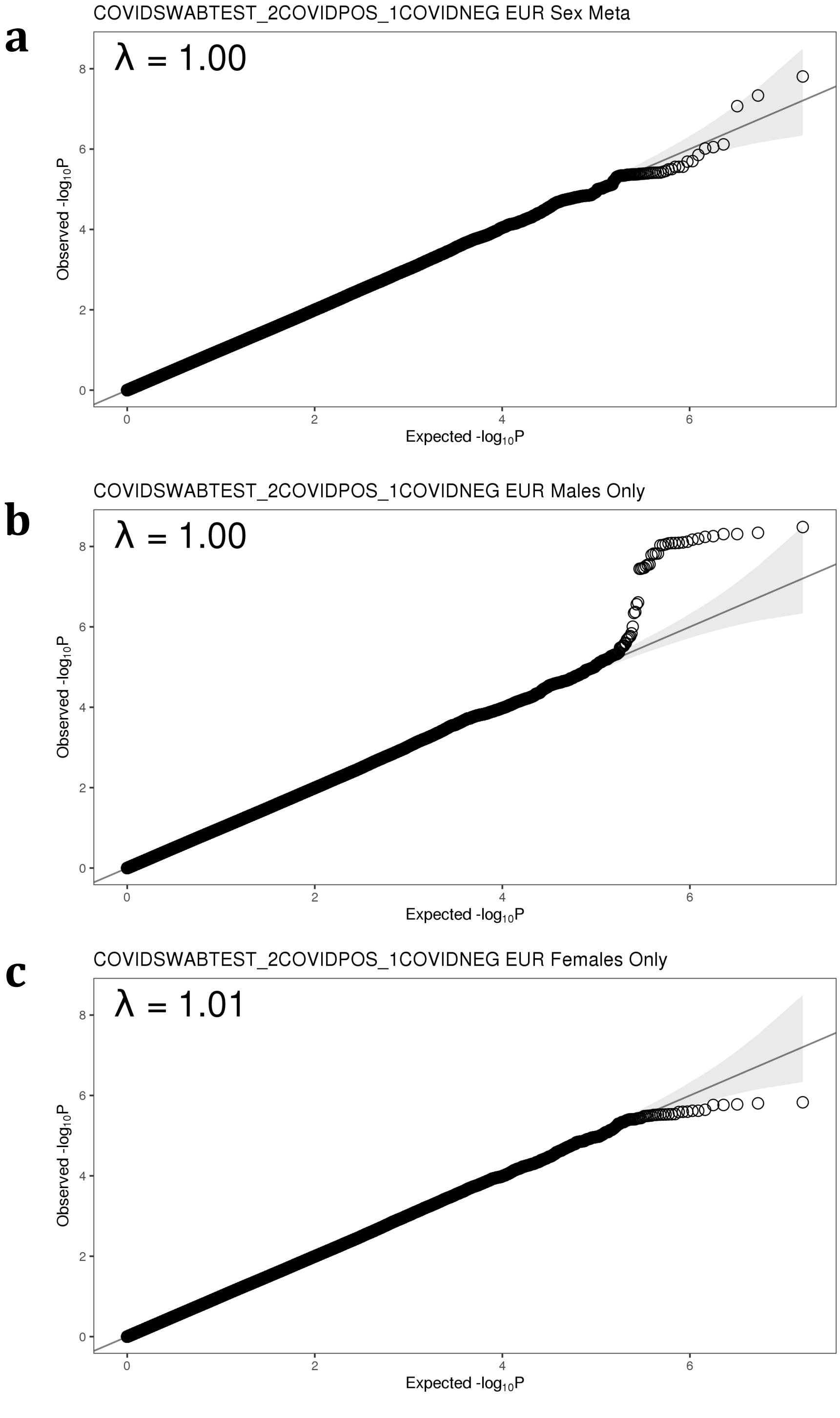


*Quantile-Quantile (QQ) plots for all variants included in the* ***(a)*** *susceptibility meta-analysis* ***(b)*** *susceptibility GWAS in males only,* ***(c)*** *susceptibility GWAS in females only. λ indicates the genomic inflation factor for each study.*

**Supplementary Figure 4:** QQplots for hospitalization GWAS

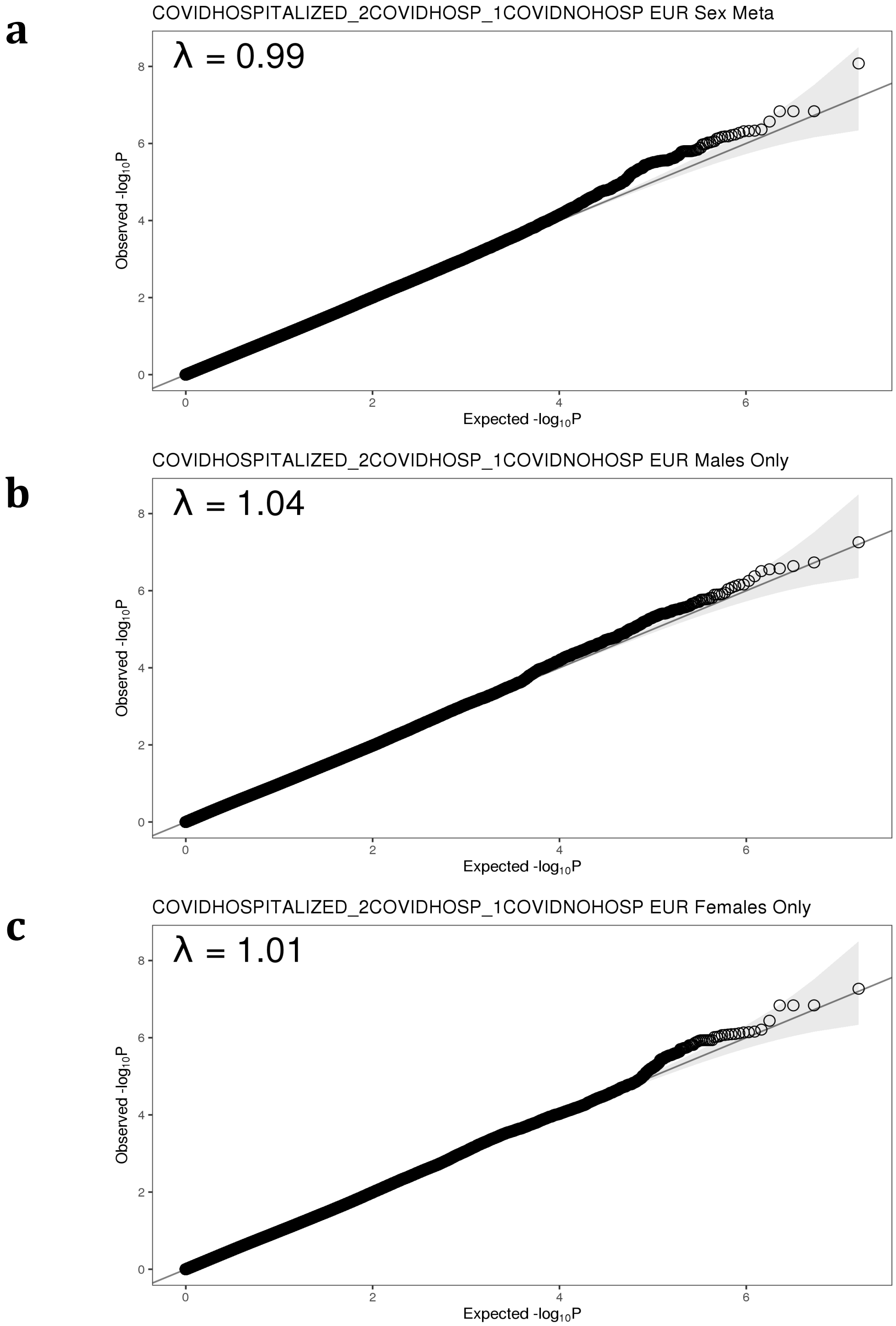


*Quantile-Quantile (QQ) plots for all variants included in the* ***(a)*** *hospitalization meta-analysis* ***(b)*** *hospitalization GWAS in males only,* ***(c)*** *hospitalization GWAS in females only. λ indicates the genomic inflation factor for each study.*

**Supplementary Figure 5:** Manhattan plots for susceptibility GWAS

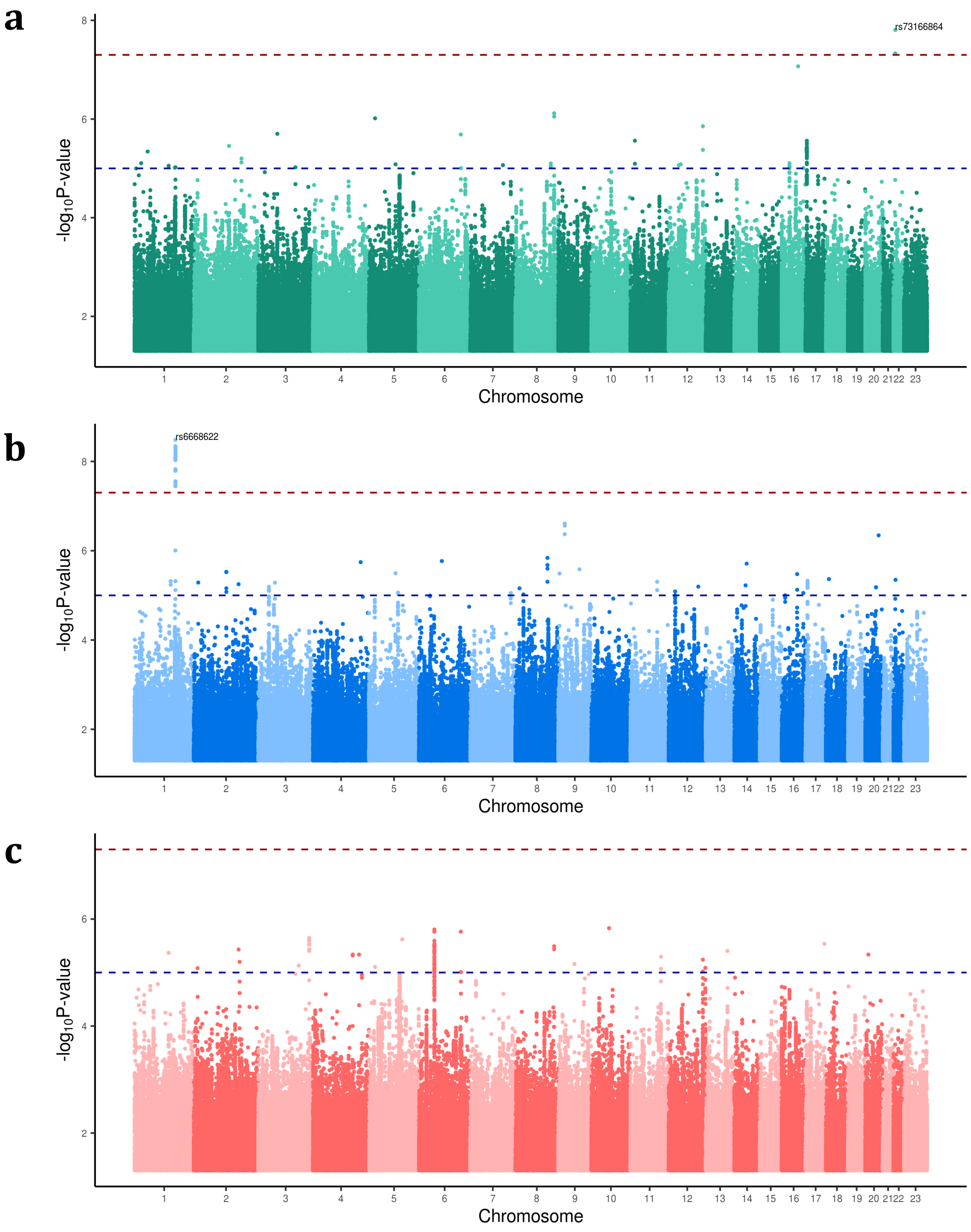


*Manhattan plots with horizontal red dashed line representing genome-wide significance (P=5x10$^{-8}$) and horizontal blue dashed line representing suggestive significance (P=1x10$^{-5}$) for the* ***(a)*** *susceptibility meta-analysis (green)* ***(b)*** *susceptibility GWAS in males only (blue),* ***(c)*** *susceptibility GWAS in females only (red). In each plot, the lead SNP dbSNP identifier for any locus surpassing P<5x10$^{-8}$ is annotated.*

**Supplementary Figure 6:** Manhattan plots for hospitalization GWAS

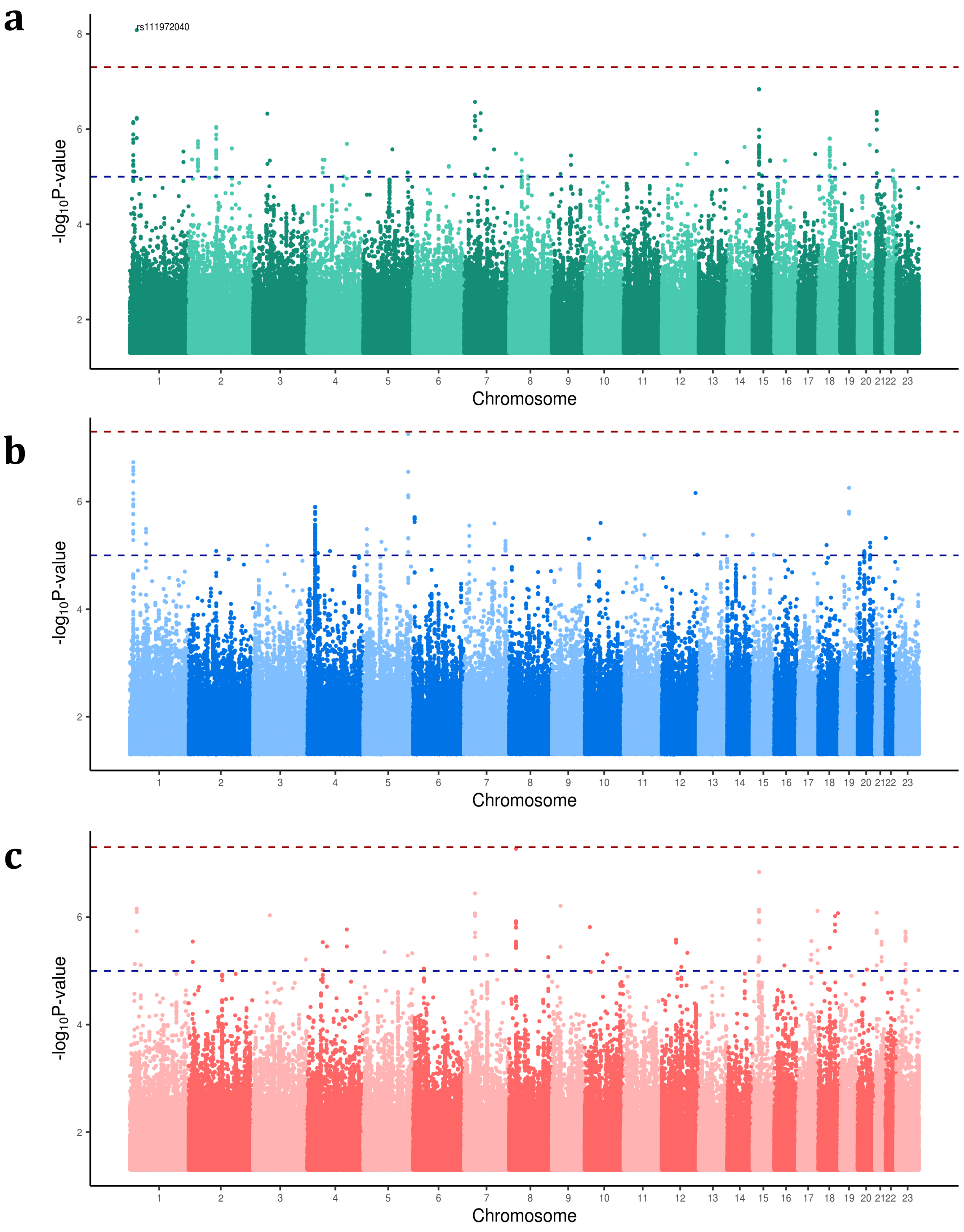


*Manhattan plots with horizontal red dashed line representing genome-wide significance ($P=5x10^{-8}$) and horizontal blue dashed line representing suggestive significance ($P=1x10^{-5}$) for the (a) hospitalization meta-analysis (green) (b) hospitalization GWAS in males only (blue), (c) hospitalization GWAS in females only (red). In each plot, the lead SNP dbSNP identifier for any locus surpassing $P<5x10^{-8}$ is annotated.*

**Supplementary Figure 7:** LocusZoom plots for *IVNS1ABP* locus

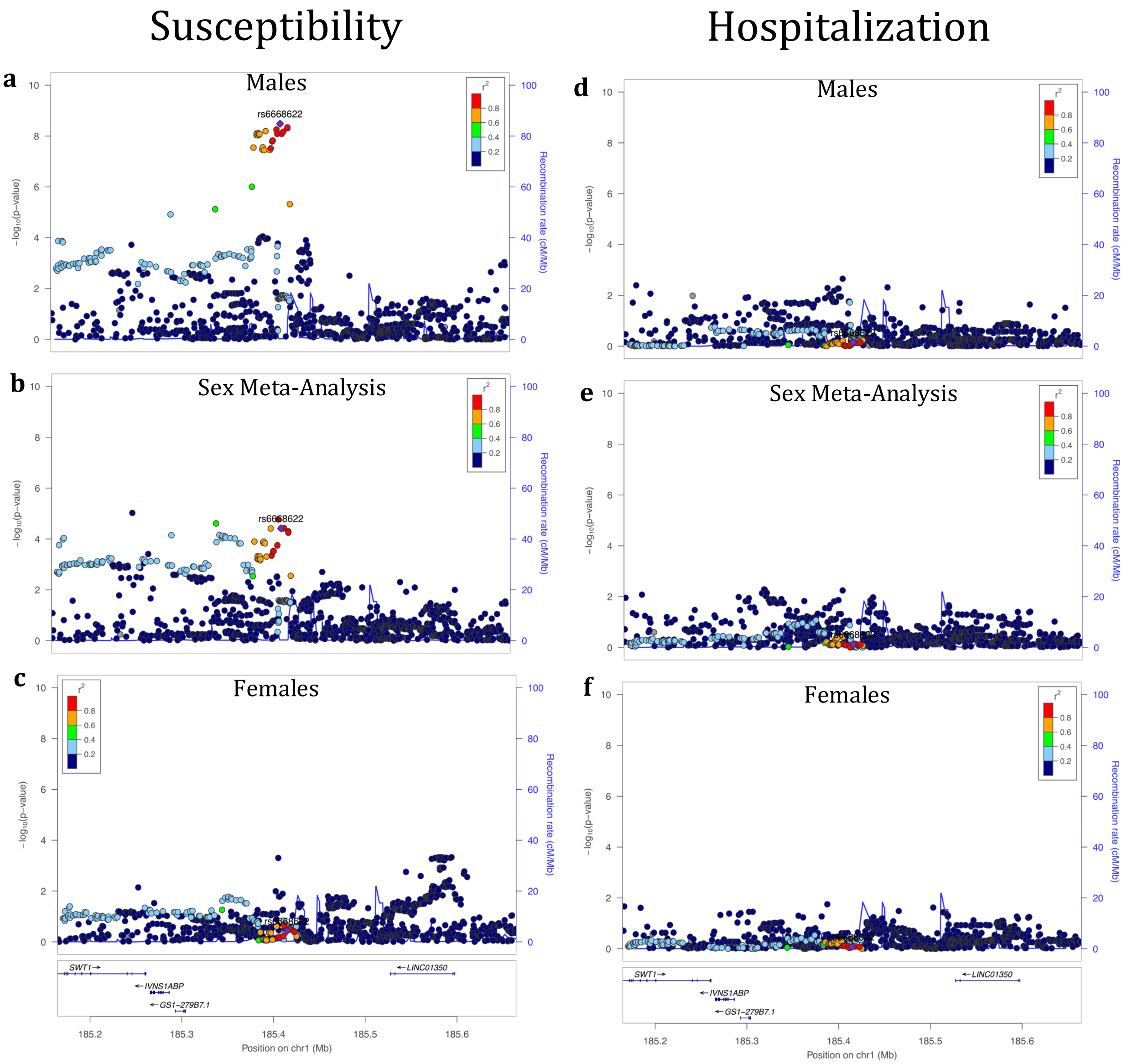


*LocusZoom plots for the IVNS1ABP locus, indexed on lead SNP rs6668622 for susceptibility GWAS in:* ***(a)*** *males only,* ***(b)*** *sex meta-analysis,* ***(c)*** *females only and for hospitalization GWAS in:* ***(d)*** *males only,* ***(e)*** *sex meta-analysis,* ***(f)*** *females only.*

**Supplementary Figure 8:** LocusZoom plots for immunoglobulin lambda locus

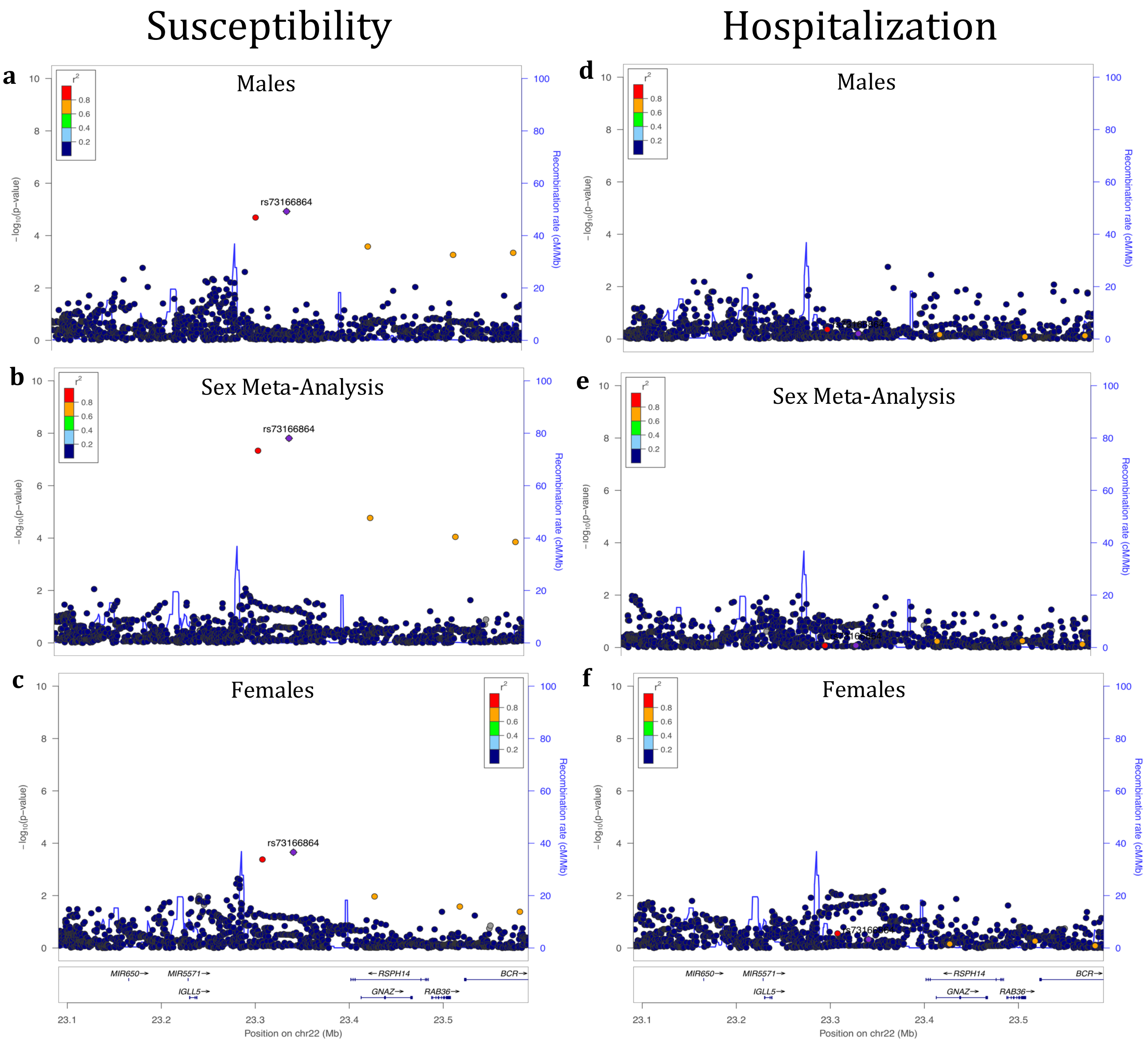


*LocusZoom plots for the immunoglobulin lambda locus, indexed on lead SNP rs73166864 for susceptibility GWAS in:* ***(a)*** *males only,* ***(b)*** *sex meta-analysis,* ***(c)*** *females only and for hospitalization GWAS in:* ***(d)*** *males only,* ***(e)*** *sex meta-analysis,* ***(f)*** *females only.*

**Supplementary Figure 9:** LocusZoom plots for *SRRM1* locus

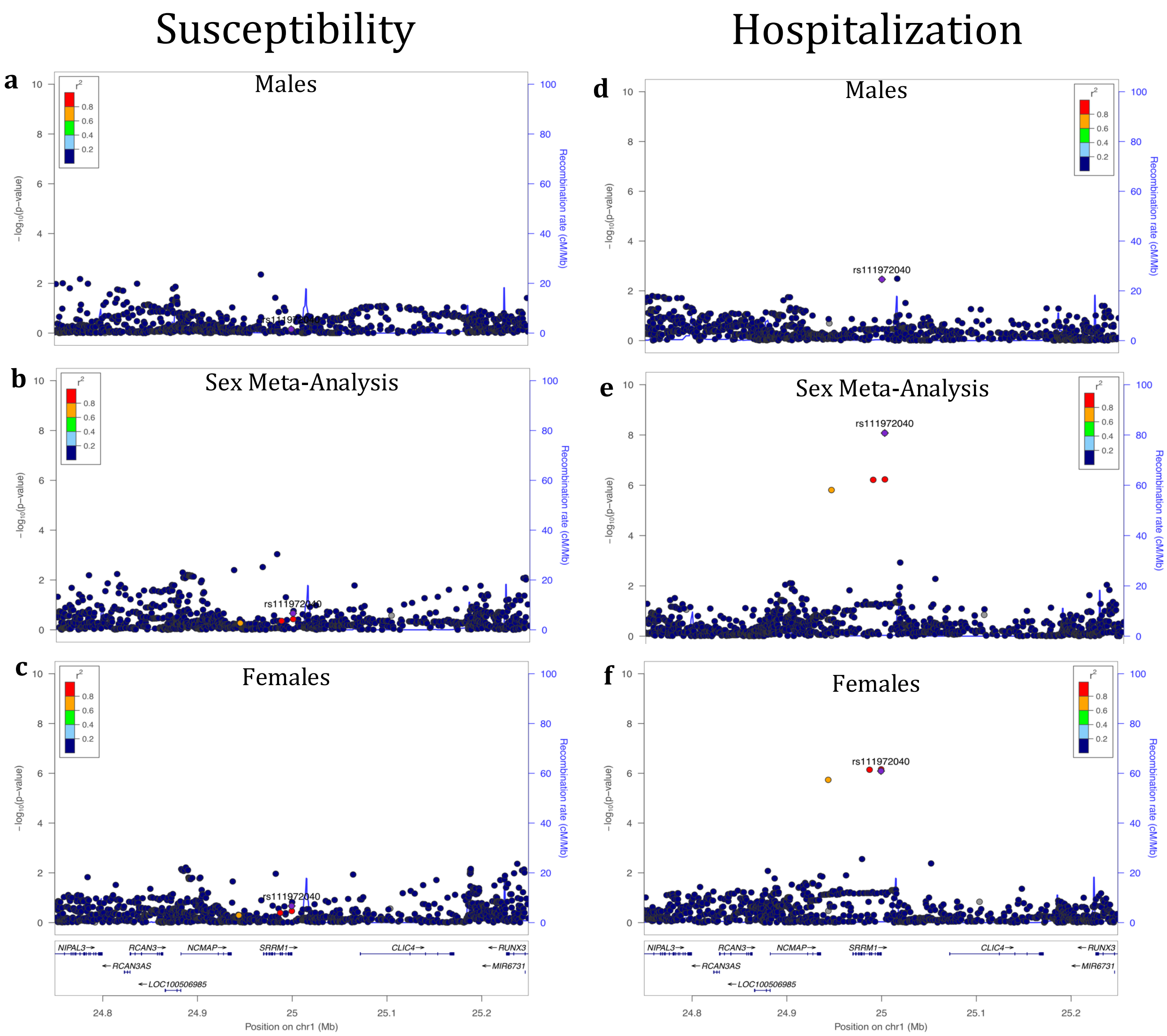


*LocusZoom plots for the SRRM1 locus, indexed on lead SNP rs111972040 for susceptibility GWAS in:* ***(a)*** *males only,* ***(b)*** *sex meta-analysis,* ***(c)*** *females only and for hospitalization GWAS in:* ***(d)*** *males only,* ***(e)*** *sex meta-analysis,* ***(f)*** *females only.*

**Supplementary Figure 10:** Principal components plot of continental ancestry groups

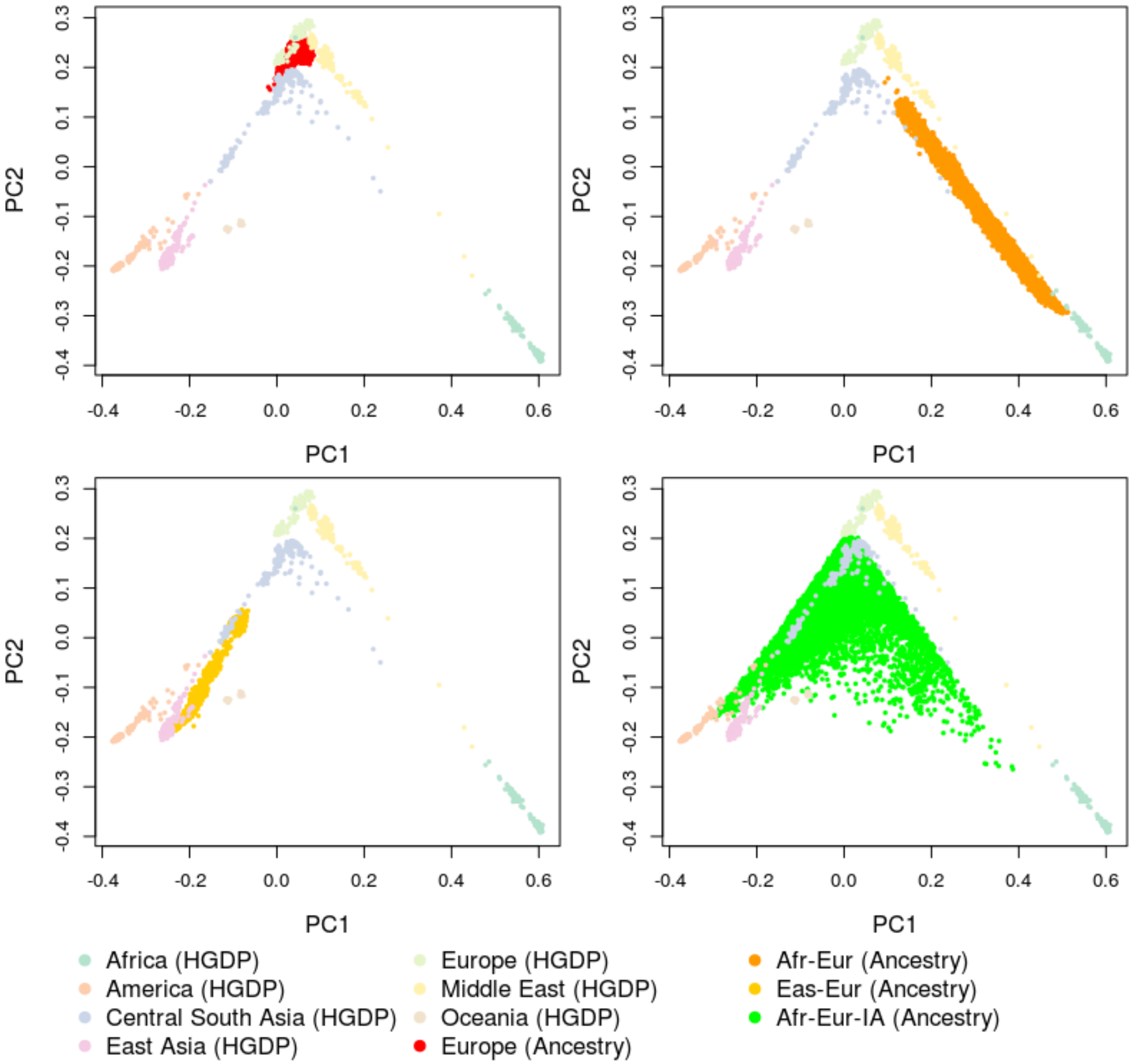


*Principal component (PC) plots of the survey cohort together with samples from the Human Genetic Diversity Project superpopulations, denoted "(HGDP)". From left to right in a clockwise direction the PC1 versus PC2 plots are shown for the following cohorts: European (Red), Admixed African-European including 100% African ancestry (Afr-Eur; Orange), Admixed East Asian-European including 100% East Asian ancestry (Eas-Eur; Dark Yellow), and Admixed Amerindian including 100% Amerindian ancestry (Afr-Eur-IA; Green).*
